# Evaluation of a novel approach to community health care delivery in Ifanadiana District, Madagascar

**DOI:** 10.1101/2020.12.11.20232611

**Authors:** Bénédicte Razafinjato, Luc Rakotonirina, Jafeta Benony Andriantahina, Laura F. Cordier, Randrianambinina Andriamihaja, Anna Rasoarivao, Mamy Andrianomenjanahary, Lanto Marovavy, Feno Hanitriniaina, Alishya Mayfield, Daniel Palazuelos, Felana Ihantamalala, Rado J.L. Rakotonanahary, Ann Miller, Andres Garchitorena, Meg G. McCarty, Matthew H. Bonds, Karen E. Finnegan

## Abstract

Despite the widespread global adoption of community health (CH) systems, there are evidence gaps in how to best deliver community-based care aligned with global best practice in remote settings where access to health care is limited and community health workers (CHWs) may be the only available providers. PIVOT partnered with the Ministry of Public Health to pilot a new two-pronged approach for care delivery in rural Madagascar: one CHW provided care at a stationary CH site while 2-5 additional CHWs provided care via proactive household visits. The pilot included professionalization of the CHW workforce (i.e. recruitment, training, financial incentive) and twice monthly supervision of CHWs. We evaluated the impact of the CH pilot on utilization and quality of integrated community case management (iCCM) in the first six months of implementation (October 2019-March 2020).

We compared utilization and proxy measures of quality of care (defined as adherence to the iCCM protocol for diagnosis, classification of disease severity, treatment) in the intervention commune and five comparison communes, using a quasi-experimental study design and relying on routinely collected programmatic data. Average per capita monthly under-five visits were 0.28 in the intervention commune and 0.22 in the comparison communes. In the intervention commune, 40.0% of visits were completed at the household via proactive care. CHWs completed all steps of the iCCM protocol in 77.8% of observed visits in the intervention commune (vs 49.5% in the comparison communes, p-value=<0.001). A two-pronged approach to CH delivery and professionalization of the CHW workforce increased utilization and demonstrated satisfactory quality of care. National stakeholders and program managers should evaluate program re-design at a local level prior to national or district-wide scale-up.

## Introduction

More than half of the world’s population lacks access to essential health services.^1^ This is especially true for low-income rural communities in Sub-Saharan Africa, where use of primary care health services decreases exponentially with geographic distance.^2^ The growing movement for universal health coverage (UHC) has been bolstered by a corresponding movement towards strengthened and professionalized community health workers (CHWs) to help address challenges with accessibility of health care.^3^ However, there is variability in the design, management and implementation of CHW programs across countries. In addition, there is limited rigorous evidence of best practices for CHW recruitment, length of training and training modalities, supervision, CHW:population ratios, and data collection and use.^4–8^

In Madagascar, a country with substantial geographic and financial barriers to care, community health workers (CHWs) provide community-based primary care services throughout the country.^9^ Ministry of Public Health (MoPH) policy dictates that there should be two CHWs in every fokontany (smallest administrative level comprising one or several villages and ranging in size from 400 to 4,500 people); CHWs are appointed by their community. CHW tasks include the delivery of integrated community case management (iCCM) for children under-five, malnutrition screening, and community health education. There is no formal education requirement for CHWs, and job-related trainings vary depending on the availability of government and partner support. CHWs often engage in other formal work and are not required to be available to provide healthcare on an established schedule. Under national policy, CHWs do not receive a salary. Instead, they sell medications for a mark-up, and thus generate income through a social marketing mechanism. CHWs are supervised monthly at a health center through group supervision by the head of the health center. Implementation of Madagascar’s community health program has often been challenged by inadequate resources, limited supervision, medication stock outs, variable training, and limited data on service provision.

In 2014, the nongovernmental organization, PIVOT, began a partnership with the Government of Madagascar to establish a model health district through integrated health system strengthening at all levels of the local public health system in Ifanadiana District. Initial interventions focused on primary care health centers and the district hospital. In 2016, PIVOT began collaborating with the MoPH to strengthen the community health program through an enhanced standard of care model which included additional training, direct supervision, modest compensation of CHWs, and support with infrastructure, equipment, and supplies.

Challenges related to recruitment, patient access, and supervision were identified. As a result, the MoPH and PIVOT initiated a new proactive community health pilot. Here, we provide an evaluation of this pilot using routinely collected program data. We describe the impact of a community health program redesign, guided by global best practices on CHW optimization, on the provision of iCCM care in one commune of rural Madagascar during the first six months of implementation.

### Setting and intervention

Ifanadiana is a rural district located in the southeast of Madagascar with a population of 182,000, nearly 33,000 of whom are children under five. The district is composed of 15 communes and 195 fokontany. In 2014, under-five mortality was 145 per 1000 live-births.^10^ A 2016 household survey found that in areas receiving PIVOT support, in the two weeks preceding the survey, 11.0% of children under-five had diarrhea, 6.8% had fever, and 11.0% had cough or difficulty breathing.^11^

In 2016, PIVOT began supporting the community health program in select communes of Ifanadiana through an enhanced standard of care model (Table 1). In 2019, guided by the principles put forth by the World Health Organization and the CHW Assessment and Improvement Matrix (AIM) Framework,^4,12^ PIVOT proposed a community health pilot in one commune of Ifanadiana District, Ranomafana. Ranomafana commune consists of eight fokontany and an estimated population of 11,960, including 2,150 children. The community health (CH) pilot is part of a strategy to achieve UHC through expanded access to high quality services.^13,14^ In partnership with the MoPH, the pilot focused on: 1) professionalization of CHWs, 2) improved care delivery through a two-pronged approach, and 3) reinforcement of the health management information system (HMIS).

**Table 1.**
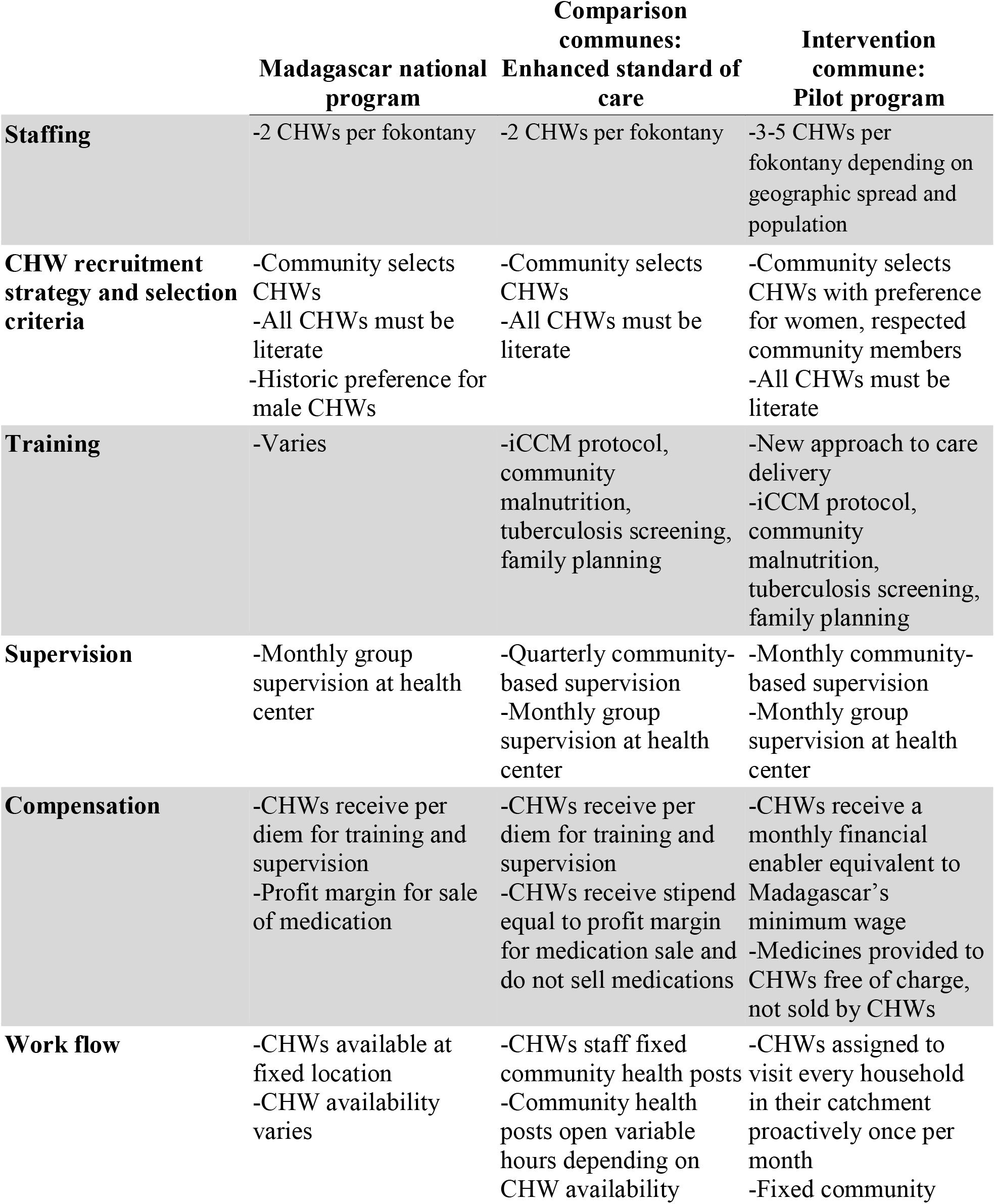
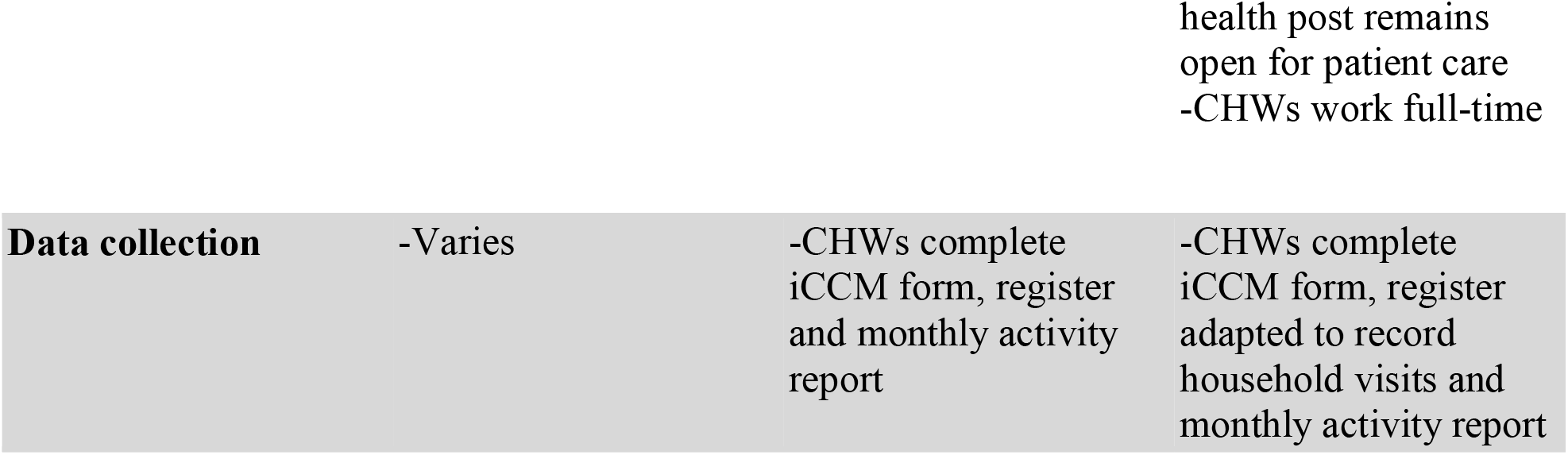
Summary of community health intervention for intervention area, control area, and the rest of the country under Madagascar’s national program.

Key components of the strategy to professionalize of CHWs included recruitment, compensation, supervision, and training (Table 1). We increased the number of CHWs in each fokontany based on population density, recruiting CHWs who were active in the community, full-time residents of the fokontany, physically capable of traveling from house to house, and with a preference for females, to address gender imbalance in the existing CHW cohort. CHWs received monthly financial motivation equivalent to Madagascar’s minimum wage and were formally evaluated every 6 months (delayed in 2020 due to COVID-19) which included a review of productivity and quality of care. A CHW could be terminated if they were not performing adequately.

PIVOT created a new cadre of community health supervisors to support the implementation of the community health pilot. Community health supervisors are health care workers with a degree in nursing or midwifery, and with experience in community health. Supervisors were trained on the iCCM protocol and then trained the newly recruited CHWs. Supervision included two activities per month: 1) community-based supervision in which the CHW and supervisor met one-on-one and the supervisor provided feedback on cases which they observed during the visit, and 2) health center-based group supervision which included discussion of activities, review of data, and short trainings on new tools or methods. During community-based visits, supervisors also provided community health education and care for sick children >5 years, whose care is not prescribed by the iCCM protocol.

Improved care delivery included a redesign of service delivery and workflow. In the pilot commune, care was delivered via a two-pronged approach: CHWs provided care at the CH site and through proactive household visits. CHWs were available five days per week, eight hours per day. One CHW in each fokontany was stationed at the CH site while other CHWs traveled on a prescribed circuit of homes. The CHW providing care at the CH site rotated regularly. CHWs in a fokontany gathered weekly to review data and discuss their activities. The redesign of care delivery was intended to overcome geographic barriers (e.g. travel distance, travel time, cost of travel) which prevent patients from seeking care from CHWs while also maintaining a functional CH site, where patients knew that they could find a CHW when one was needed. During household visits, the CHWs actively sought out sick children and followed up on children previously diagnosed with malaria, diarrhea, pneumonia, or malnutrition. CHWs completed a follow up visit with sick children three days after diagnosis to determine if symptoms had resolved and, for severe cases where referral was required, CHWs visited the next day to ensure that the sick child had visited the health center.

Strengthening the HIMS included support in the development and use of new management and data collection tools. During field-based visits, supervisors reviewed data collection processes and form completion to ensure that CHWs were collecting high quality data on their activities. At monthly health-center based supervision, CHWs reviewed and submitted monthly aggregate activity reports.

### Program evaluation

We used data routinely collected as part of the CH program to evaluate the impact of the pilot. We extracted data on the number of children seen at community health sites and via proactive home visits from monthly CHW activity reports to measure CHW utilization by children under-five. We extracted information on adherence to the iCCM protocol, a proxy measure for quality of care, from this same monthly report. The monthly HMIS report provides a limited number of indicators on iCCM protocol adherence for all CHW visits. We extracted measures of correct treatment of diarrhea (child with diagnosis of simple diarrhea provided with ORS), respiratory infection (child with diagnosis of suspected pneumonia treated with amoxicillin), malaria (child with malaria diagnosed by rapid diagnostic test treated with ACT), and speed of fever treatment (child with any fever seen by CHW within 24 hours of fever onset). We also obtained data on CHW adherence to the iCCM protocol from the observation checklist completed by CHW supervisors during field-based supervision visits. The observation checklist provides data on protocol adherence for a subset of the CHW visits completed each month (see Appendix for a French language version of the observation checklist). Supervision visits were conducted at both the community health site and during proactive home visits. Using data from the observation checklist, we calculated a summary measure of quality of care, correct care. Correct care measures how many of the total steps of the iCCM protocol the CHW completed correctly based on the child’s diagnosis and includes components of diagnosis, disease classification, treatment, and counseling of the caregiver on follow up. We calculated correct care following the initiation of the pilot as a revised supervision tool was introduced as part of the pilot.

We used a quasi-experimental study design to determine the impact of the CH pilot. We compared outcomes in one intervention commune, Ranomafana, before and after the implementation of the pilot program with those in five other communes of Ifanadiana. These other communes were supported by PIVOT and the MoPH under the enhanced standard of care model as defined in Table 1; this support has been rolled out over time (Appendix 1). The pilot was implemented from October 2019 to March 2020. We assessed utilization and program delivery in the intervention and comparison communes before and after the start of the pilot. We used a two-sample t-test to compare continuous outcomes and a chi-squared test for categorical outcomes. We used a t-test to compare correct care, which is a percentage of steps completed corrected correctly with a score which ranges from 0% to 100%. Data analysis was completed in R version 3.5.2.

This study was approved by the Secretary General of the Ministry of Public Health of Madagascar and was determined to be not human subjects research by Harvard Medical School’s Institutional Review Board.

## Findings

### Professionalization of CHWs

The total number of CHWs across the 8 fokontany of the intervention commune increased from 16 to 28, an average of one CHW per 427 people. CHWs per fokontany ranged from 3 to 5. There were 108 CHWs across the five comparison communes, an average of 21.6 per commune and one CHW per 532 people (SD: 165.1).

Supervisors completed 154 supervision visits in the intervention area and directly observed 281 sick child visits to document quality of care and provide feedback. Nearly all (91.7%) of CHWs were supervised twice per month during the six-month pilot in the intervention commune. CHWs were observed providing care to an average of 1.6 (SD: 0.9) children per month. In the comparison communes, supervisors completed 363 community visits. Approximately half (46.3%) of CHWs were supervised twice per month and 95.6% of CHWs were supervised at least once per month during group supervision.

### Care delivery

During the six-month pilot period, CHWs in the intervention commune completed 3,628 visits with children under-five, representing a 283.5% increase in visits from the year prior. Average monthly per-capita utilization was 0.28 or 3.4 visits per capita per year (Figure 1). Average monthly per capita utilization was 0.22 in the comparison group for the same six-month period. Monthly per capita utilization increased over time and with each level of enhancement of support, although these increases were not statistically significant. Per capita utilization increased over time across all eight fokontany in the intervention commune (Figure 2).

**Figure 1.**
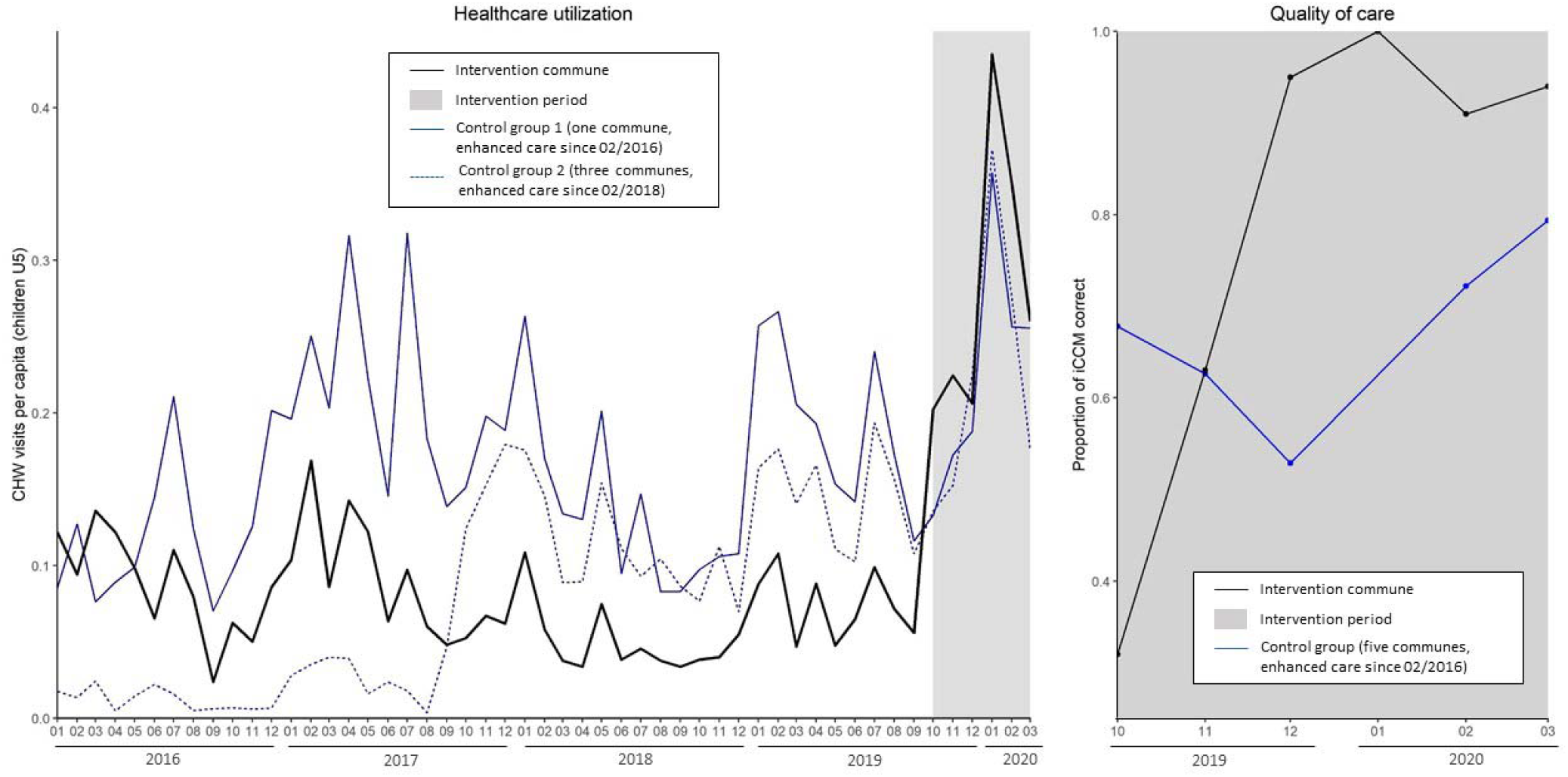
Average monthly per capita utilization of CHWs (left) under by intervention group from January 2016 to March 2020. A comparison of correct care (right), measured through direct observation of the CHW during an iCCM, in the intervention commune (black) and comparison communes receiving enhanced standard of care (blue) from October 2019 to March 2020.

**Figure 2.**
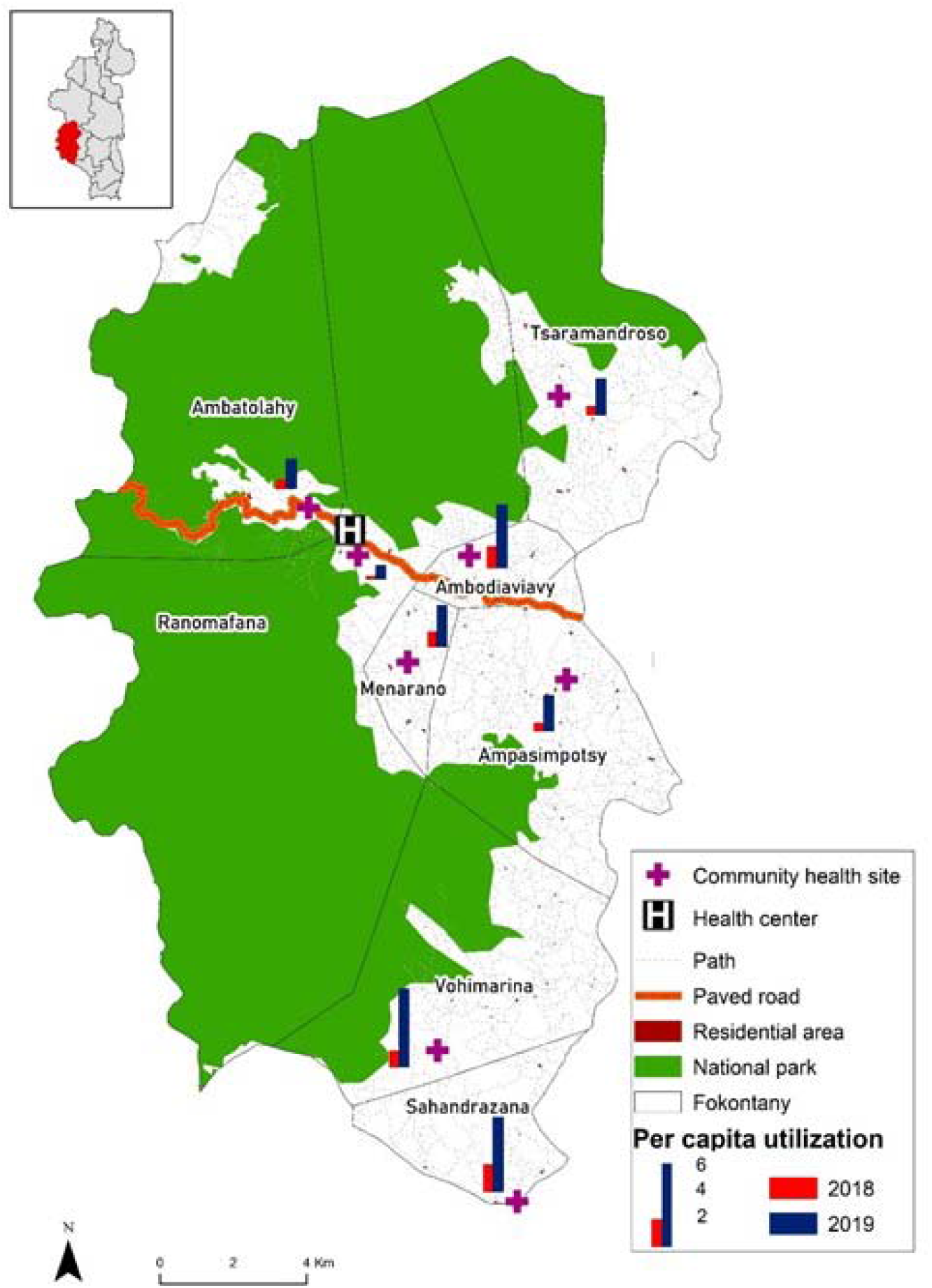
Per capita community health utilization by fokontany in the intervention commune in October 2018-March 2019 (red) and October 2019-March 2020 (blue). The district of Ifanadiana is in the inset with the intervention commune in red.

In the intervention commune, 40.0% of CHW visits were via proactive care at the household (Figure 3); on average, 79.4% of households were visited at least once every month. CHWs in the intervention commune evaluated 2591 cases of fever (71.4% of visits), 435 cases of simple diarrhea (12.0% of visits), 1529 cases of pneumonia (42.1% of visits), and 556 cases of cough or cold (15.3%) during the six-month period; children may be diagnosed with more than one illness during a visit. Less than half (40.0%) of fever cases had a positive malaria rapid diagnostic test. In the comparison communes, CHWs completed a total of 13,239 visits with children under-five during the six-month study period. Of these visits, 86.0% were for fever, 9.4% for simple diarrhea, 42.0% with pneumonia, and 11.9% with cough or cold. Among fever cases, 49.3% were positive for malaria using a rapid diagnostic test. There were significant differences (p-value<0.001) in the rates of diagnosis among sic children in the intervention and comparison communes for the six-month period for malaria, diarrhea, and cough or cold, but not for pneumonia.

**Figure 3.**
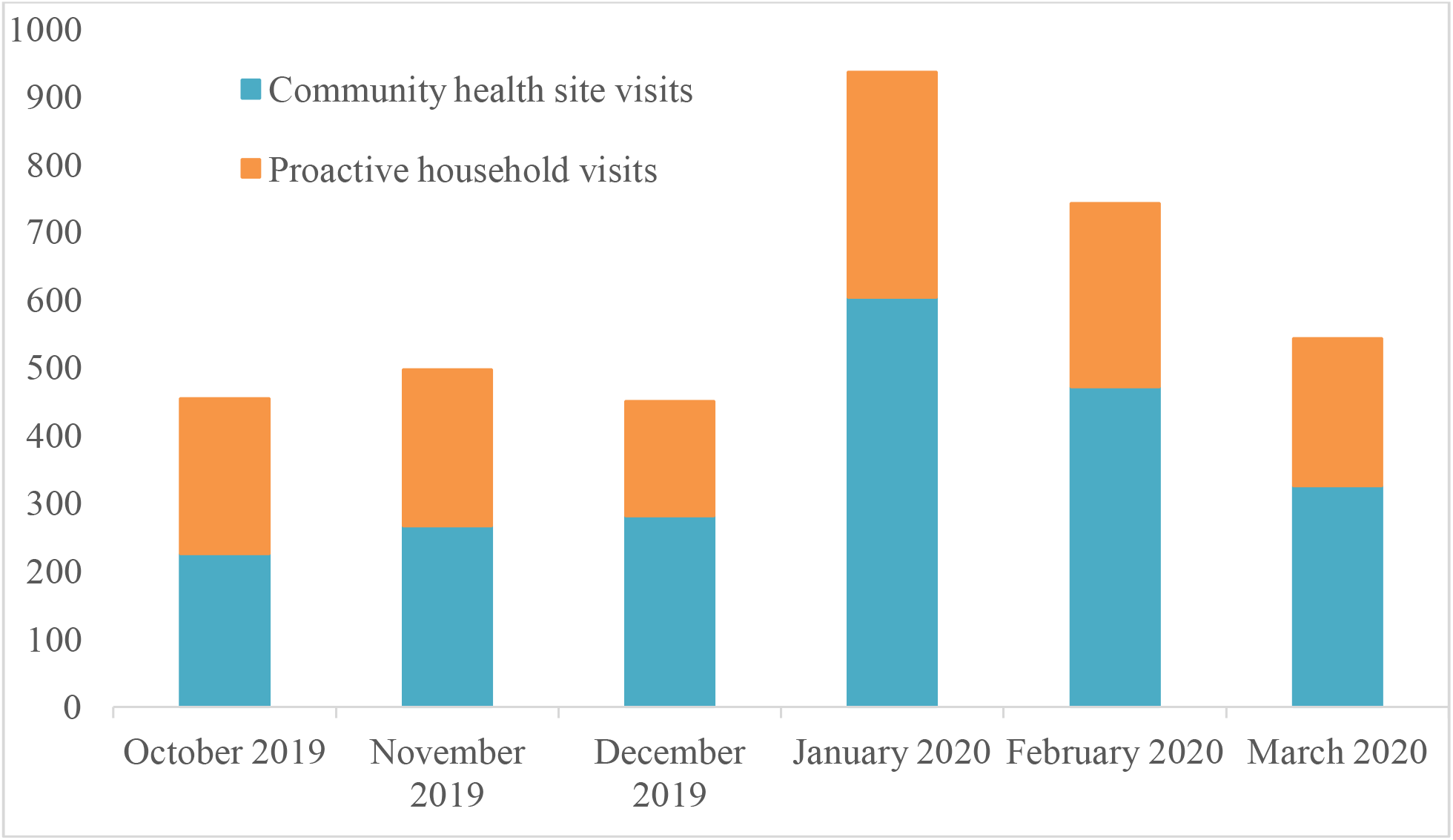
CHW visits by location in the intervention commune during the six-month implementation of the CH pilot. Household visits are in (blue) and CH site visits are in (orange).

Correct care, as measured through supervisor observation, increased from 32.0% in October 2019 to 94.0% in March 2020 in the intervention commune. Over the six-month study period, 77.8% of children received care consistent with all aspects of the iCCM protocol in the intervention commune; in the comparison communes, CHWs demonstrated complete protocol adherence in 50.2% of observed visits (p-value of difference <0.001) (Table 2). Likewise, CHWs in the intervention commune demonstrated better evaluation of danger signs (96.4% vs 90.2% in comparison area, p-value<0.001), correct treatment of illness (96.7% vs 90.4%, p-value=0.001), and counseling of caregiver on treatment and disease management (84.1% vs 67.4%, p-value<0.001). CHWs in intervention and comparison communes demonstrated similarly high rates of correct diagnosis of illness. While quality of care remained relatively constant in the comparison communes, all measures of quality improved in the intervention area from the first to last month of the study period. The greatest improvement was observed in counseling of the caregiver (October=45.7%, March=95.3%, p-value<0.01) and referral to health center (October=55.6%, March=100.0%, p-value=0.04) which nearly doubled over the study period. By the end of the six-month pilot, the quality of care provided by newly recruited CHWs according to the iCCM protocol was equal to that of existing CHWs who had previously been working in community health (see Appendix).

**Table 2.** Quality of care of iCCM provided by CHWs during observed visits from October 2019-March 2020.

|  | Intervention commune | Comparison communes | p-value |
| --- | --- | --- | --- |
| Number of clinical encounters observed | 281 | 821 |  |
| Children correctly cared for according to iCCM protocol | 77.8% | 50.2% | <0.001 |
| Correct evaluation of danger signs | 96.4% | 90.2% | <0.001 |
| Correct diagnosis of illness | 99.4% | 98.7% | 0.18 |
| Correct treatment of illness | 96.7% | 90.4% | 0.001 |
| Caregiver counseled | 84.1% | 67.4% | <0.001 |
| Child correctly referred to health center | 90.2% | 75.6% | 0.01 |

Disease-specific quality of care measures were variable over time (Figure 4). During the six-month pilot period, the intervention commune provided higher rates of treatment of malaria (intervention=79.3%, comparison=57.1%, p-value=0.04), pneumonia (intervention=65.0%, comparison=56.3%, p-value=0.14), and diarrhea (intervention=88.3%, comparison=69.3%, p-value<0.001). Only 10.3% of fever cases were seen within 24 hours of symptom onset in the intervention commune, compared to 12.9% in the comparison communes during the six-month study period.

**Figure 4.**
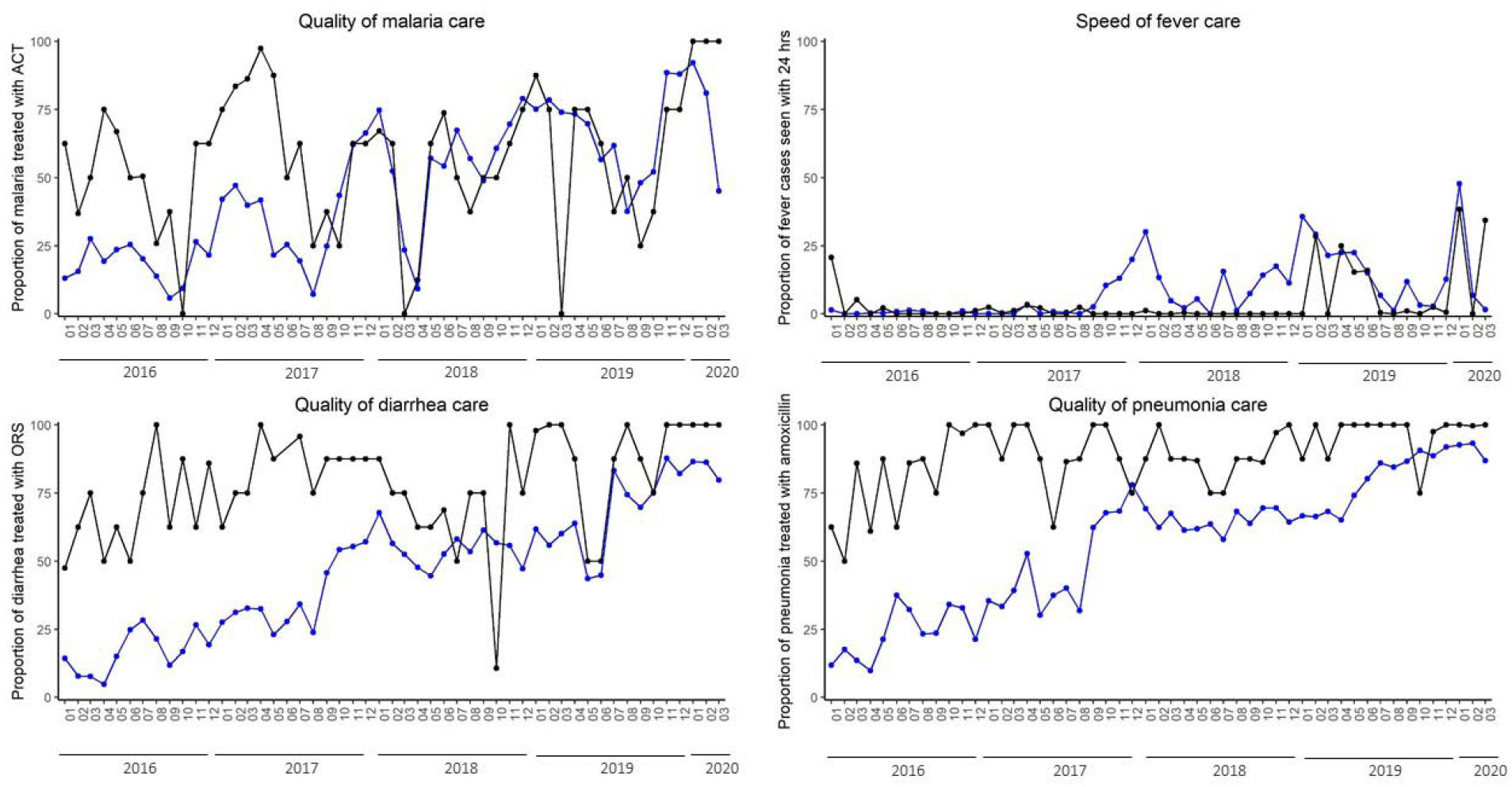
Disease-specific quality of care measures from iCCM patient visits for malaria treatment with ACT (top left), speed of fever treatment (top right), diarrhea treatment with ORS (bottom left), and pneumonia treatment with amoxicilin (bottom right) comparing the intervention commune (black) with the average of the quality measure from the five comparison communes (blue).

### HMIS

In both intervention and comparison communes, the monthly CHW activity report was submitted on time and without missing data. Over the study period, the concordance rate between the iCCM form (used to record detailed data during the patient visit), iCCM register (a line listing summary of each patient visit), and the monthly report (an aggregate summary of the activity of all CHWs in a commune) was 88.7% in the intervention commune and 80.1% in comparison areas. Higher rates of concordance indicate data which is consistent across sources.

## Discussion

A two-pronged approach to the delivery of iCCM by CHWs, including both proactive and health post-based care, accompanied by professionalization of the cadre, and HMIS support, was implemented in rural Madagascar and demonstrated improvements in service delivery and quality of care. When compared to other communes implementing an enhanced standard of care, the pilot program equaled or exceeded the comparison communes on measures of utilization and quality of care. Evaluation of the pilot program provides lessons for national stakeholders and program managers on the impact of program re-redesign and reveals important considerations for scale-up.

The CH pilot led to moderate increases in service delivery and improved quality of care. Over the six-month study period, almost half of visits completed by CHWs in the intervention commune were proactive household visits. The increase in per capita utilization during the intervention period suggests that the two-pronged approach to care increased access. This is further supported by the increase in per capita utilization in fokontany which are farthest from the health center. However, a key component of the iCCM protocol is referral to a health facility when a child presents with danger signs. Given limitations in data collection, we were unable to determine if patients sought care for treatment at the health facility after being referred by a CHW. We were able to determine that referral rates were high in both intervention and comparison areas.

The pilot program introduced professionalized CHWs to the CH system. The pilot required intensive program management and placed high expectations for productivity on CHWs and supervisors. To ensure that CHWs were supervised twice per month, supervisors from other communes were called to the intervention area to provide community-based supervision. Although effective in meeting supervision targets and improving quality of care, such intensive intervention may not be sustainable. Supervision often requires multiple days of travel on foot for supervisors to reach CHWs in remote communities. Under national guidelines, supervision of CHWs is the responsibility of health center managers; the development of a cadre of CHW supervisors is one innovation of the pilot project and recognizes that existing health center staff do not have sufficient time to travel to supervise all affiliated CHWs as they provide care. The World Health Organization names supportive supervision as an important component of CH programs and research highlights the importance of supervision in establishing effective high-quality care, although evidence on the impact of supervision on quality of care is mixed.^6,15^ Although the development of a cadre of dedicated supervisors was important for the launch of the community health pilot, for long term implementation and scale up, PIVOT will recommend identification and training of high-performing CHWs to serve as peer mentors and perform many of the functions of supervisors.

When studying Madagascar’s national community health program, Brunie et al found that CHWs reported high levels of satisfaction with their work, but also high levels of financial uncertainty and most relied on subsistence farming for their livelihoods.^16^ As part of the pilot, CHWs received a financial incentive equivalent to Madagascar’s minimum wage (approximately $70 USD) per month, which helped alleviate some of these financial concerns. Sustainability and harmonization are key factors when considering CHW salary as part of iCCM programs in low resource countries.^17^ This pilot provides evidence of the feasibility of providing remuneration as part of program management; a more rigorous costing analysis is needed to determine financial feasibility of national scale-up.

This rapid evaluation comprised the first six months of the pilot program and was intended to generate evidence on the impact of a program re-design prior to adoption in other areas of the district. The findings were shared with the MoPH, which approved the pilot, to provide information on different modes of care delivery for the country’s rural districts. The CH program is an important component of Madagascar’s UHC plan.

We generated early evidence of the impact of the program; additional evaluations are needed to determine if short-term gains in utilization and quality of care are sustained. Our assessment of the pilot intervention would be enhanced by an understanding CHW and community perceptions of the program.

The intervention commune demonstrated significant improvements in utilization and quality of care, while other communes maintained fairly high rates of both under an enhanced standard of care. We noted variability in quality of care by disease, which merits further research to understand which factors drive differences in quality between and among CHWs. The study period concluded shortly after the first cases of COVID-19 were diagnosed in Madagascar and no cases were identified in Ifanadiana District during the study period. The Ministry of Health and PIVOT’s clinical teams are developing guidelines to modify CHW protocols to provide high quality care and reduce the transmission of COVID-19. The impact of COVID-19 on care-seeking and program design in rural communities is not yet well understood.

There are pressing questions about how CHW programs should be designed and implemented. Global evidence is mixed and often weak.^15^ Making use of available data collected as part of PIVOT’s integrated data platform, our rapid evaluation of a two-pronged pilot program in a rural commune in Madagascar can contribute to global evidence on the optimal design of community programs. Moreover, this experience can provide actionable lessons for Madagascar’s national community health program on program re-design to align with global best practice on community health program principles.

## Data Availability

Data remain the property of the Madagascar Ministry of Public Health. If interested in accessing the data, please contact the corresponding author who will help facilitate access.

## Acknowledgements

We would like to thank the health system actors who helped with the conception, implementation and realization of community health activities: the health center staff, the district health team, the Ministry of Health, the supervisors, and especially the CHWs and communities of Ifanadiana District.

## Notes

### Competing Interest Statement

BR, LR, LFC, AR, MA, LM, FM, AM, DP, FI, RJLR, AM, MHB, and KEF receive salary support from PIVOT. JBA and RA are employees of the Ministry of Public Health in Madagascar.

### Funding Statement

This work was supported by funding provided by Conservation, Food and Health Foundation; CRI Foundation; IZUMI Foundation; MJS Foundation; Preston-Werner Ventures; Wagner Foundation. The funders played no role in study design, manuscript preparation, statistical analysis, or the decision to publish.

### Author Declarations

Madagascar's Ministry of Public Health approved this study. The Institutional Review Board of Harvard Medical School approved the study and determined that it was not human subjects research.

